# Reacting to outbreaks at neighboring localities

**DOI:** 10.1101/2020.04.24.20078808

**Authors:** Ceyhun Eksin, Martial Ndeffo-Mbah, Joshua S. Weitz

## Abstract

We study the dynamics of epidemics in a networked metapopulation model. In each subpopulation, representing a locality, the disease propagates according to a modified susceptible-exposed-infected-recovered (SEIR) dynamics. In the modified SEIR dynamics, individuals reduce their number of contacts as a function of the weighted sum of cumulative number of cases within the locality and in neighboring localities. We consider a scenario with two localities where disease originates in one locality and is exported to the neighboring locality via travel of exposed (latently infected) individuals. We establish a lower bound on the outbreak size at the origin as a function of the speed of spread. Using the lower bound on the outbreak size at the origin, we establish an upper bound on the outbreak size at the importing locality as a function of the speed of spread and the level of preparedness for the low mobility regime. We evaluate the critical levels of preparedness that stop the disease from spreading at the importing locality. Finally, we show how the benefit of preparedness diminishes under high mobility rates. Our results highlight the importance of preparedness at localities where cases are beginning to rise such that localities can help stop local outbreaks when they respond to the severity of outbreaks in neighboring localities.

## 1. Introduction

Early detection of disease outbreaks at their location of origin provide a chance for local containment and time to prepare in other locations. Such preparation may enable locations connected to the origin to become more aware of the outbreak and develop a stronger response to the disease especially when it is not contained. The success of containment strategies is highly dependent on the ability of promptly detecting most infectious individuals in a given location. The recent outbreak of the COVID-19 virus has shown that successful containment efforts are highly challenging when many latently infected and asymptomatic but infectious individuals can travel undetected between locations [1, 2, 3, 4].

In the ongoing COVID-19 outbreak, localities in the US are continuing to see alarming surges in the number of cases and hospitalized individuals at different times, driven in part by differences in introduction and lift-off of the epidemic in local communities [5, 6]. Reducing mobility between localities can delay the overall epidemic progression. However, prior research suggests that the final outbreak size is not strongly affected by travel restrictions unless combined with a strong reduction in transmission within the locality [2, 7, 8, 9]. In the US, local authorities have implemented non-pharmaceutical interventions, e.g., declaring emergency or issuing stay at home orders, at different times. Community response to these interventions differ across localities [10, 11, 12, 13, 14, 15, 16]. Hence, there is growing concern that mismatched timing of response efforts could lead to a failure of containment [17].

Here we develop a simplified model to assess the combined effects of mobility, local response to disease prevalence, and the level of alertness prior to disease surge in a locality. To do so, we consider a networked-metapopulation model [18, 19, 20, 21, 22]. Prior metapopulation models have focused on the heterogeneity of the populations [21], hierarchical connectivity structures among populations [18], and local responses of populations [22]. These models are fitted to various childhood diseases [18], e.g., measles, pertussis, as well as, the common cold [22] and SARS [23]. More recently, the effects of local lockdowns on the COVID-19 outbreak have been investigated using metapopulation models [15, 10]. Here, we assume the disease progresses according to susceptible-exposed-infected-recovered (SEIR) dynamics within each population or locality (similar to [24]). Within each population susceptible individuals can become exposed via contact with infected individuals in the same locality. SEIR models are a standard approach to model epidemiological dynamics including pandemic influenza [25] and COVID-19 [26, 27].

In the present context, we extend SEIR models to include the effects of behavior changes on local disease progression. We assume individuals change their behavior and reduce their contacts proportional to disease prevalence, i.e., the ratio of infected and recovered [28, 29]. In addition, behavior in a locality can be influenced by the disease prevalence in neighboring localities. That is, we introduce awareness-driven social distancing models that account for interaction between localities not just in terms of the flow of individuals, but also in terms of the flow of information that leads to raised awareness (social distancing and preparation). While prior works considered local social distancing efforts determined by local disease prevalence [22, 15, 10], we include a mechanistic model of the influence of the outbreak sizes at neighboring localities. Our aim is to quantify the combined effects of inter-locality mobility, and behavior changes in response to local and external disease prevalence. As we show, behavior changes driven by the awareness of outbreaks in neighboring localities can reduce the spread of a newly imported disease in connected populations.

## 2. Methods

We consider a networked metapopulation model of epidemic dynamics. At each population, the disease propagates according to SEIR dynamics given a homogeneous population. In addition, we assume there is constant travel in and out of each population. The flow of travelers constitute only healthy (susceptible) individuals, and those that are latently infected (exposed). The dynamics at locality *i* are given as follows:

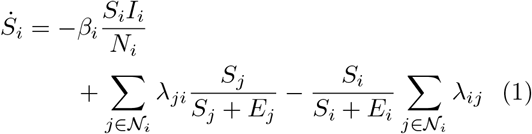

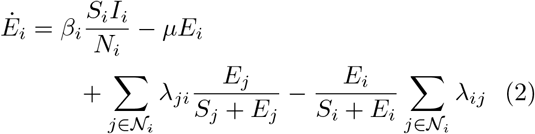

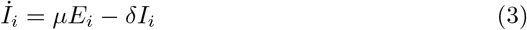

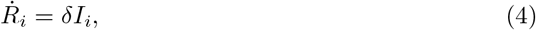

where *β*_*i*_ is the transmission rate at location *i, λ*_*ij*_ is the flow of individuals from location *i* to neighboring location *j, µ* denotes transition rate from exposed (pre-symptomatic) to infected (symptomatic), and *δ* is the recovery rate. We denote the neighboring localities of *i* with *N*_*i*_. We assume total flow in and out of a location are equal, i.e., *λ*_*ij*_ = *λ*_*ji*_. The total mobility flow from *i* to *j* include susceptible and exposed individuals proportional to their size in the population. We assume infected individuals are successfully detected, and thus cannot travel between localities. The model does not include mobility of recovered individuals. Mobility of recovered individuals may reduce the outbreak in localities as they may serve as barriers and reduce the outbreak [30]. Here, we neglect possible barrier effects of recovered mobility individuals in order to focus on the effects of awareness-based social distancing.

The transmission rate at location *i* depends on the inherent infection rate *β*_0_ and social distancing due to cumulative disease prevalence,

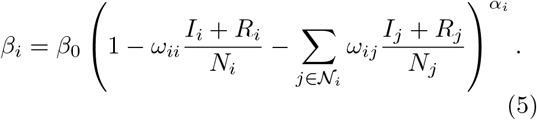

In the social distancing model, individuals reduce their interaction with others proportional to the ratio of cumulative cases, defined as the ratio of infectious and recovered in the population, at locality *i* and neighboring localities of *i* [28, 29, 31]. Here, we consider social distancing in a broader sense as the impact of all individual and public health measures that reduce social contact between individuals (e.g. in the case of COVID-19 this may include six feet physical distancing, restriction on social and economic activities, and partial lock downs). The term inside the parentheses is the awareness at locality *i* caused by disease prevalence. The weight constant *ω*_*ii*_ ∈ [0, 1] determines the importance of disease prevalence at locality *i* versus the importance of disease prevalence at neighboring *𝒩*_*i*_ localities, *ω*_*ij*_ ∈ [0, 1]. We assume the weights sum to one, i.e., 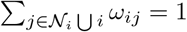. The exponent constant *α*_*i*_ represents the strength of response to the disease awareness. It determines the overall distancing at locality *i* based on the awareness. If *α*_*i*_ = 0, there is no distancing response to the awareness at locality *i*. Note that the awareness term inside the parentheses is always less than or equal to 1. Thus, the larger *α*_*i*_ is, the larger is the distancing response at locality *i* to disease prevalence. We refer to the case with *α*_*i*_ = 1 as the linear distancing model.

In the following, we consider two localities with equal population sizes *N*_1_ = *N*_2_, unless otherwise stated. The disease starts at locality 1 with 0.1% of the population in exposed state, and spreads over to locality 2 via undetected exposed individuals traveling from 1 to 2. The travel between localities does not change the population sizes, i.e., we assume *λ*_12_ =*λ*_21_. We set 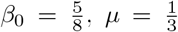, and 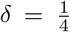 based on the rates estimated at [1] for the COVID-19 outbreak in China. The reproduction number at locality *i* is 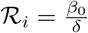 = 2.5 for *i* ∈ {1, 2}. Note that the standard SEIR model is recovered when *α*_*i*_ = 0 and *λ*_*ij*_ = 0 for all localities.

## 3. Results

### 3.1. Mobility and Social Distancing

As a baseline we consider no distancing response, i.e., *α*_*i*_ = 0 for all *i* = {1, 2} (Figure 1). We find that Locality 2 follows an almost identical disease trajectory as Locality 1 approximately 38 days after Locality 1 when *λ*_*ij*_ = 0.01%. The difference in peak times of the two localities increases from 10 days to 51 days as *λ*_*ij*_ decreases from 1% to 0.001% (Figure 1). Moreover, as the mobility rate increases, the outbreak at Locality 1 becomes larger than the outbreak at Locality 2 while Locality 1 experiences a lower peak than Locality 2—see Figure 1(Left). The intuition is that the duration of the epidemic in Locality 1 is longer due to the large number infected individuals traveling from Locality 2 after the peak time of Locality 1. This difference in outbreak sizes is negligible compared to the effects of social distancing.

**Figure 1:**
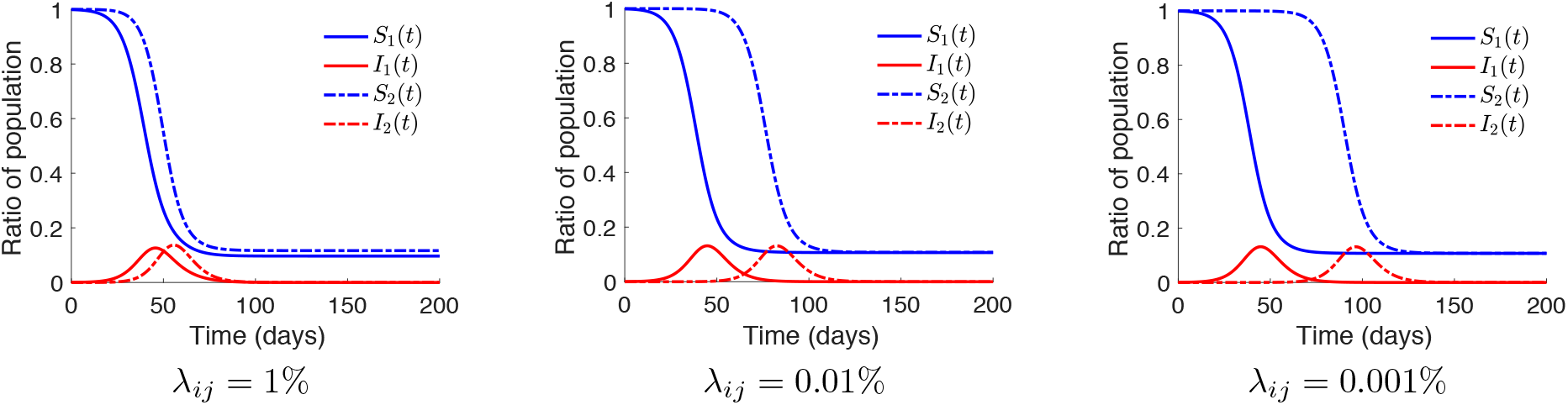
Networked SEIR model with no-distancing. Two localities are connected with travel rates *λ*_*ij*_ ∈ {1%, 0.01%, 0.001%}. The disease propagates in both localities according to SEIR dynamics with no response to disease prevalence, i.e., *α*_*i*_ = 0. Blue and red lines show the ratio of susceptible and infected individuals in a locality, respectively. The differences in time of peaks are 10, 38, 51 days respectively for *λ*_*ij*_ ∈ {1%, 0.01%, 0.001%}. Final outbreak sizes of localities 1 and 2 are almost identical for low mobility regimes.

Next, we consider the effect of social distancing. For this, we assume localities only put weight on disease prevalence at their own locality, i.e., *ω*_*ii*_ = 1 for *i* ∈ {1, 2}. Figure 2 shows the percentage reduction in final outbreak size and peak ratio of infected at Locality 2 as localities become more responsive, i.e., as *α*_*i*_ increases. When the distancing is linear *α*_*i*_ = 1, the reductions in peak and outbreak size are near 25%. Reduction in both metrics reaches 70% when *α*_*i*_ = 5. This range of values of the impact of social distancing on disease transmission is consistent with empirical estimates for COVID-19 in Europe and the US [32, 33]. While outbreak size continues to decrease with *α*_*i*_ increasing, there does not exist a critical threshold of *α*_*i*_ that stops the disease spread in a locality. The failure to stop an outbreak with awareness is due to the proportionality of the social distancing to the cumulative number of cases [28].

**Figure 2:**
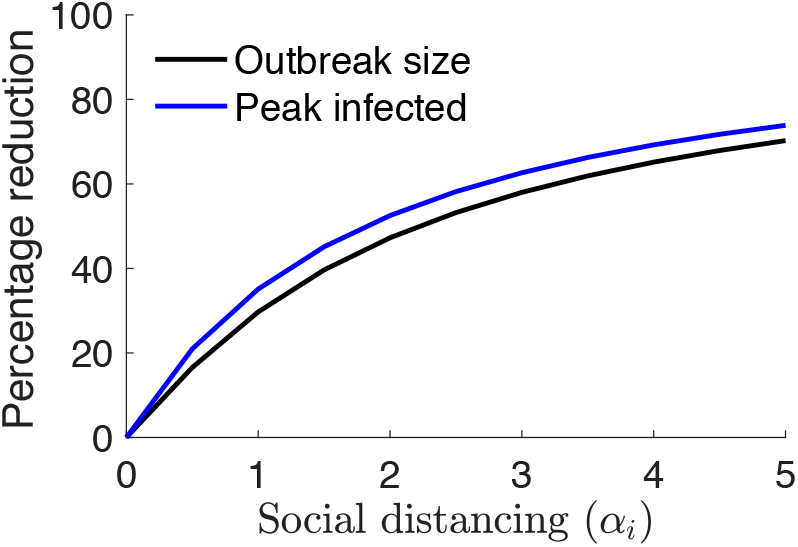
Percentage reduction in outbreak size and ratio of infected at peak with respect to increasing social distancing exponent (*α*_*i*_). We measure the reduction with respect to the no-distancing case (*α*_*i*_ = 0). In both cases, the mobility per day is *λ*_12_ = *λ*_21_ = 0.001% of the population.

### 3.2. A lower bound on the outbreak size at the origin

Final size relationships for SEIR dynamics without mobility connect the strength of an epidemic (reproduction number) to the number of individuals not infected at the end of the epidemic *S*(∞). In the present case, such relationships constitute an analogous lower bound for the outbreak size at the origin in a scenario without mobility (*λ*_*ij*_ = 0) and *ω*_12_ = 0,

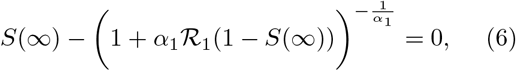

where 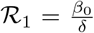 is the reproduction number at the origin. Above, we assume compartments (*S, E, I, R*) in the model dynamics represent the fraction of population in the corresponding stage of the disease. In obtaining the relation in (6), we consider a modified social distancing model in which we also include fraction of exposed *E* in the distancing term—see Appendix A. When individuals reduce their interactions proportional to the cumulative number of exposed cases, the social distancing is stronger than (5). Thus, the solution to (6) for *S*(∞) is an *upper bound* for the fraction of final susceptible individuals, which means it is a *lower bound* for the fraction of final recovered individuals

(*R*(∞)). For the linear distancing case *α*_1_ = 1, we obtain the closed form solution to (6),

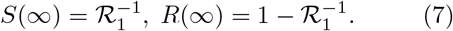

Figure 3 compares the actual outbreak sizes with the upper bound for *S*(∞) obtained by solving (6) for different values of *α*_1_. We observe that the upper bound solution, denoted with 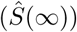, is loose by a constant amount that is approximately equal to 0.04 in both *k* = 1 and *k* = 3. We also note that this upper bound provides a good approximation of the outbreak size at Locality 2 when mobility is low and *ω*_21_ = 0.

**Figure 3:**
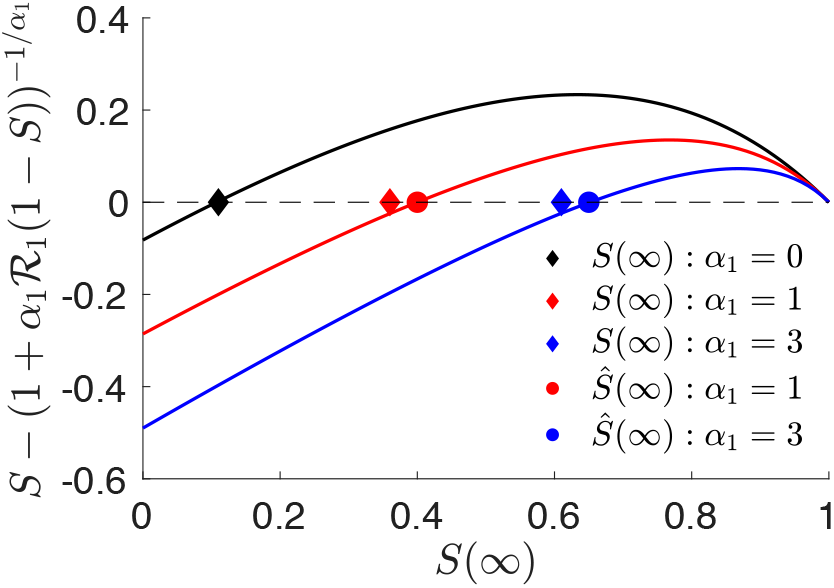
Upper bound values of *S*(∞) obtained by solving (6) for *k* = 1 and *k* = 3. We let ℛ_1_ = 2.5. Lines correspond to the left hand side of (6). Circle dots show the solution to (6). Diamond dots are *S*(∞) values obtained by simulating the SEIR model in (1)-(4) with *β*_*i*_ given in (5). For *k* = 0, we use standard speed-size relations for the SEIR model without social distancing [34]. Note that the relation for the standard SEIR model is exact. Thus, diamond and circle dots overlap for *k* = 0. The difference between the upper bound for *S*(∞) 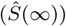 and the simulated *S*(∞) is relatively constant for different values of *α*_1_.

### 3.3. Adopted awareness

We analyze the effect of awareness at Locality 2 caused by the outbreak in Locality 1. We denote the weight *ω*_21_ associated with this awareness as the adopted awareness weight. We assume Locality 1’s awareness is not shaped by the outbreak at Locality 2, i.e., *ω*_11_ = 1. In this scenario, the adopted awareness should be interpreted as individuals in Locality 2 reducing contacts, e.g., practice social distancing, based on the awareness created by the outbreak at Locality 1. When the disease starts in one location (Locality 1) and moves to a neighboring locality (Locality 2) via travel of exposed, the adopted awareness distancing term at Locality 2 is a measure of its *preparedness*.

We begin by using the lower bound for the outbreak size at the origin 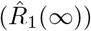 to obtain an upper bound for the outbreak size at Locality 2 as a function of *ω*_21_. There exists a time *T* > 0 such that for all *t*> *T*, we have 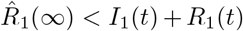 where 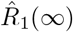 is obtained by solving (6) for 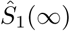 and setting 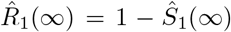. Consider the social distancing model *β*_2_(*t*) in (5). For *t*> *T*, we have

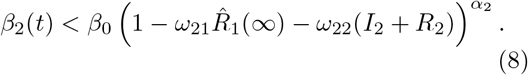

As mobility slows down, i.e., *λ*_12_ → 0, the threshold time *T* approaches zero as well. Thus, in the slow mobility regime, the inequality above holds for almost all times. By ignoring the social distancing based on local awareness, we obtain the following upper bound on the infection rate at the importing locality (Locality 2),

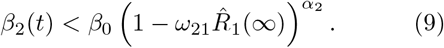

Note that the right hand side is a constant that depends on the lower bound of the outbreak size at the origin and the strength of response at Locality 2 (*α*_2_). Given the constant upper bound in (9), we use the speed-outbreak size relation for the standard SEIR model, e.g., see [34, 35], to obtain a lower bound for *S*_2_(∞) at neighboring locality,

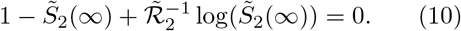

where we define

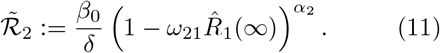

The solution to (10) given by 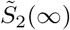 provides a lower bound for *S*_2_(∞) in the SEIR model with social distancing (1)-(5). Note that we obtain the speed-outbreak size relation for the standard SEIR model when *α*_2_ = 0. In Appendix B, we demonstrate how the solution to (10) changes as a function of strength of responses at localities—see Figures S1-S2.

We compare the outbreak size at Locality 2 from simulating (1)-(5) with the upper bound obtained by solving (10) for a range of adopted awareness values *ω*_21_ ∈ [0, 1] (Figure 4). We observe that the upper bound is loose when the adopted awareness is close to zero. This is reasonable since in deriving the bound we removed the social distancing with respect to local disease prevalence. The accuracy of the upper bound improves as the adopted awareness constant increases. Indeed, as per our assumptions, as *λ*_21_ → 0, the upper bound would tend to the actual outbreak size when *ω*_21_ = 1.

**Figure 4:**
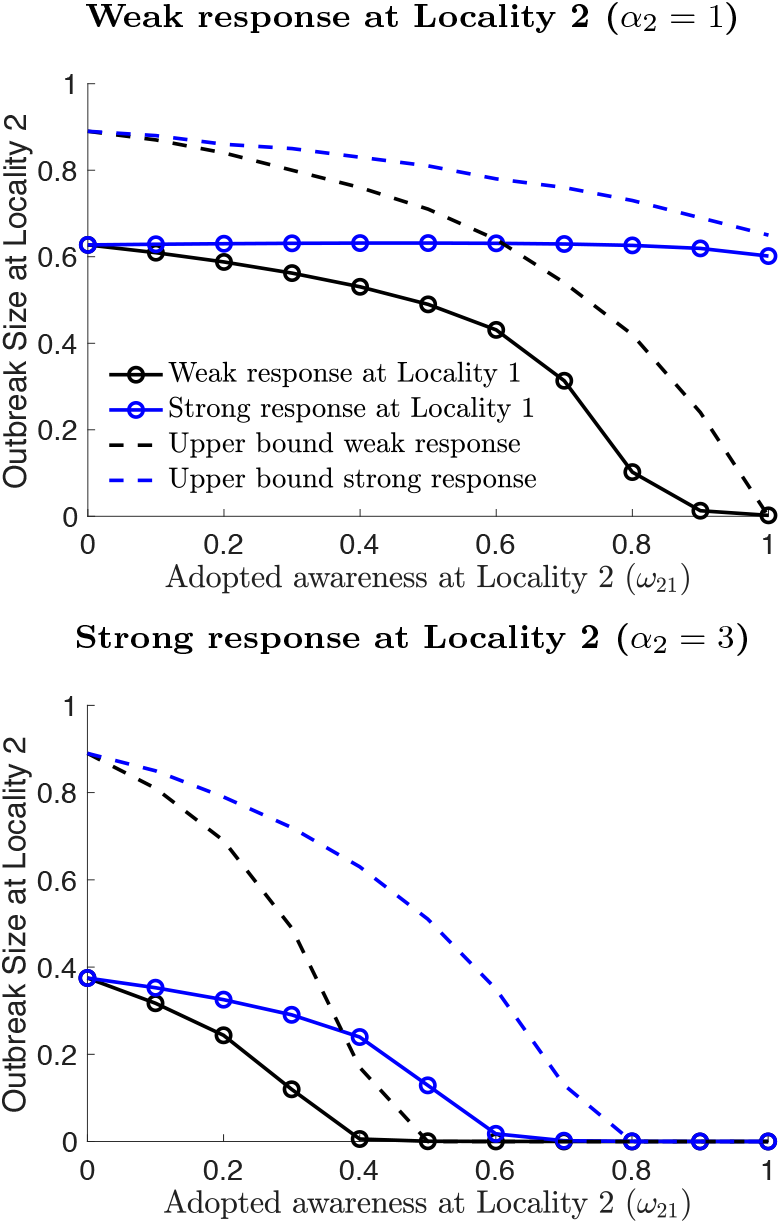
Outbreak size at Locality 2 with respect to adopted awareness *ω*_21_. (Top) Weak (*α*_2_ = 1) and (Bottom) Strong (*α*_2_ = 3) responses at Locality 2. Mobility is set to *λ* = 0.001%. Weak and strong responses at Locality 1 correspond to *α*_1_ = 1 (black) and *α*_1_ = 3 (blue), respectively. The outbreak size at Locality 2 decreases with increasing adopted awareness values. The decrease is sharper when response at the origin is weak. Corresponding theoretical upper bound values (shown by dashed lines) are tighter at larger adopted awareness values. In the Bottom figure, thecritical threshold values 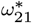 above which disease does not propagate approximately equal to 0.5 and 0.8 respectively for weak (black) and strong (blue) responses at the origin.

Both the outbreak size at Locality 2 and the associated upper bound monotonically decrease as adopted awareness (*ω*_21_) increases for the given strengths of response at the origin *α*_1_ ∈ {1, 3}. This means Locality 2 is better off reacting to the outbreak at Locality 1, as this will lead to an early strong response to the disease. Indeed, the decrease of the outbreak size at Locality 2 with respect to the adopted awareness constant is faster when the response at Locality 1 is weak—compare blue and black lines within Top and Bottom panels in Figure 4. The reason for this is that a weaker response at Locality 1 results in a higher ratio of cumulative cases, which means higher awareness at Locality 2. Going in the other direction, if the strength of response at Locality 1 further increases (*α*_1_ > 3), it is possible that increasing adopted awareness increases the outbreak size at Locality 2. This means the monotonic decrease in the outbreak size at Locality 2 with respect to increasing adopted awareness is contingent on the strength of response at Locality 1 and the mobility constants.

The preparedness at Locality 2 can result in stopping the outbreak from spreading at Locality 2. Indeed, we observe in Figure 4 (Bottom) that there exists a critical threshold for the adopted awareness constant *ω*_21_ > 0.4 above which outbreak size is near zero for Locality 2.

Next, we use the upper bound for the outbreak size at Locality 2 to compute an upper bound for the critical threshold value of the adopted awareness constant given *α*_1_ and *α*_2_ values. In order to obtain this threshold, we rely on the result that when *ℛ*_2_ < 1, the disease will die out in a standard SEIR model. Note that 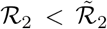 with 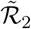 is as defined in (11). Thus, the disease will not spread at Locality 2 if 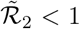. Solving this condition for *ω*_21_, we get the following threshold

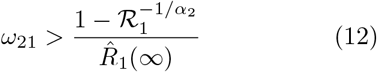

where 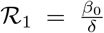 is the reproduction number and 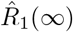 is the lower bound on the outbreak size at the origin obtained by solving (6). From (12), we see that the critical threshold value for adopted awareness 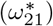 increases with increasing *α*_1_ and decreases with increasing *α*_2_. In Figure 4 (Bottom), we see that the theoretical critical threshold values are close to the actual (simulated) *ω*_21_ values above which the disease does not propagate in Locality 2.

For *α*_1_ = 1, we obtain a close form solution for
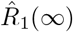 in (7), which yields

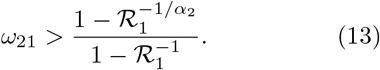

The threshold value above is an increasing function of *α*_2_. That is, Locality 2 can avoid an outbreak with a smaller adopted awareness constant (*ω*_21_) as *α*_2_ increases. For *α*_2_ = 1, we have the right hand side equal to 1. This means there does not exist a level of preparedness, i.e., a value of *ω*_21_ ∈ [0, 1], such that the disease is eliminated at Locality 2. This confirms the results shown in Figure 4 (Top)— see solid and dashed black lines decreasing toward 0 as *ω*_21_ goes to 1. At *ω*_21_ = 1, the adopted awareness is equal to the right hand side of (13) where the prediction is that the disease can still spread in Locality 2.

### 3.4. Effect of adopted awareness on total outbreak size

While the above analyses show that Locality 2 can benefit from a heightened awareness due to a weak response at Locality 1, this awareness is a direct result of the lack of control at Locality 1. Indeed, a larger outbreak at Locality 1 yields a lower outbreak size at Locality 2. Here, we address conditions in which the reduction in the outbreak size at Locality 2 due to the increase in the outbreak size at Locality 1 is larger than the increase in the outbreak size at Locality 1.

We begin by focusing on the total outbreak size defined as the sum of outbreak sizes in both localities, i.e., *R*_1_(∞)+*R*_2_ (∞), as a global measure of the effects of adopted awareness. We find that when the response at Locality 2 is weak (*α*_2_ = 1), there exists a level of preparedness (*ω*_21_ ≈ 0.7) above which the total outbreak size is smaller when the response at Locality 1 is weak—see blue line dip below the black line around *ω*_21_ ≈ 0.7 in Figure 5 (Top). In contrast, when the response at Locality 2 is strong, there does not exist an adopted awareness constant value where a weak response at Locality 1 is better than a strong response at Locality 1 in terms of total outbreak size—see Figure 5 (Bottom). Lastly, the total outbreak size is always lower when the response at Locality 2 is strong—compare Figures 5 (Top) and (Bottom). These observations indicate that we obtain the best outcome in terms of total outbreak size when both localities respond strongly, and Locality 2 has an adopted awareness constant value above the critical threshold value.

**Figure 5:**
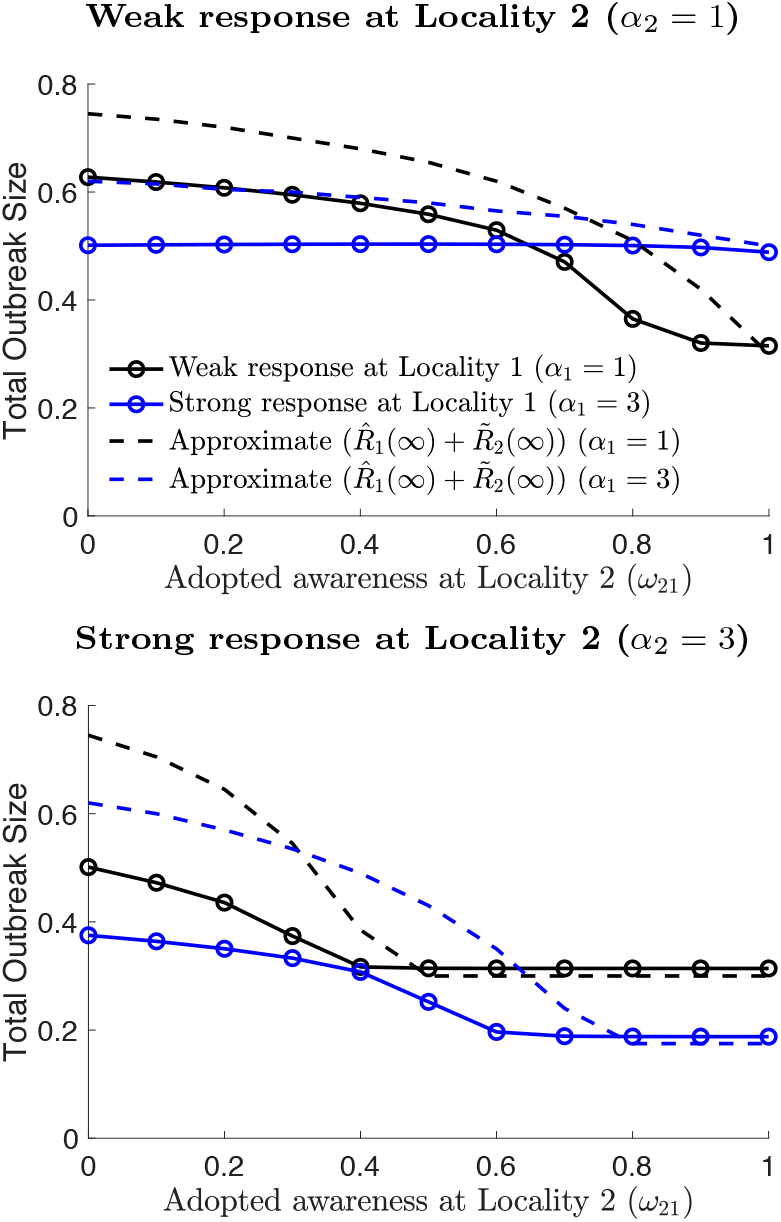
Total of outbreak sizes at localities 1 and 2 with respect to adopted awareness *ω*_21_. (Top) Weak (*ω α*_2_ = 1) and (Bottom) strong (*α*_2_ = 3) responses at Locality 2. Mobility is set to *λ* = 0.001%. Weak and strong response at Locality 1 correspond to (*α*_1_ = 1) and (*α*_1_ = 3), respectively. There exists a critical adopted awareness constant value in Top where the total outbreak size is lower in the scenario where both localities respond weakly compared to the scenario where Locality 1 has a strong response. The critical value for the adopted awareness constant value can be found by looking at the intersection of the solid black line with the solid blue line for the corresponding mobility value. When both localities respond strongly to the disease in Bottom figure, such a critical adopted awareness constant value does not exist.

In Figure 5, we also provide a theoretical approximation of the total outbreak size computed by adding the upper bound for the outbreak size in the origin 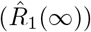 to the lower bound for the outbreak size at Locality 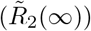. This total 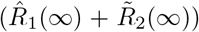 is neither an upper bound nor a lower bound. We see that the approximation error is mostly dominated by the error in the upper bound 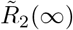 for small values of the adopted awareness constant. The approximation is a lower bound of the total outbreak size for all adopted awareness values above the critical adopted awareness constant computed using (12)—see dashed lines in Figure 5(Bottom) lying below the corresponding solid lines.

### 3.5. Effects of mobility rates

Thus far, we have focused our analysis on the effects of adopted awareness given a slow mobility regime (*λ*_12_ = 0.001%). As per the discussion in Section 3.1, the outbreak times between localities get closer as mobility increases. Given higher mobility rates, the cumulative number of cases at Locality 1 will be lower by the time disease begins to spread at Locality 2. Thus, we expect the benefit of adopted awareness at Locality 2 to be lower with increasing mobility.

We measure the benefit of adopted awareness (*ω*_21_) by comparing the outbreak size at Locality 2 given a positive adopted awareness value *ω*_21_ > 0 with the outbreak size when adopted awareness constant is zero, i.e., *ω*_21_ = 0. Following the discussion above, given a positive adopted awareness constant value *ω*_21_ > 0, the potential benefit of adopted awareness reduces as mobility increases (Figure 6). We see that the decrease in the benefit of preparedness is slow up until a mobility rate value. After a certain mobility value *λ*_12_ ≈ 0.05%, the decrease in the benefit of adopted awareness is sharper. Regardless, we observe that when the response at the origin is weak, it is better to have a higher level of adopted awareness—see Figure 6 (Top). The magnitude of benefits of adopted awareness is reduced when the response at the origin is strong—see Figure 6 (Bottom). Indeed, lower adopted awareness values can yield smaller outbreak sizes at Locality 2 for high mobility—observe that the benefit value dips below zero at higher mobility rates in Figure 6 (Bottom). The reason for the negative benefit is that the disease severity at Locality 2 quickly exceeds the outbreak size at Locality 1 when the response at the origin is strong at high mobility rates, which means that Locality 2 is better off social distancing based on the local outbreak size rather than the outbreak size at Locality 1.

**Figure 6:**
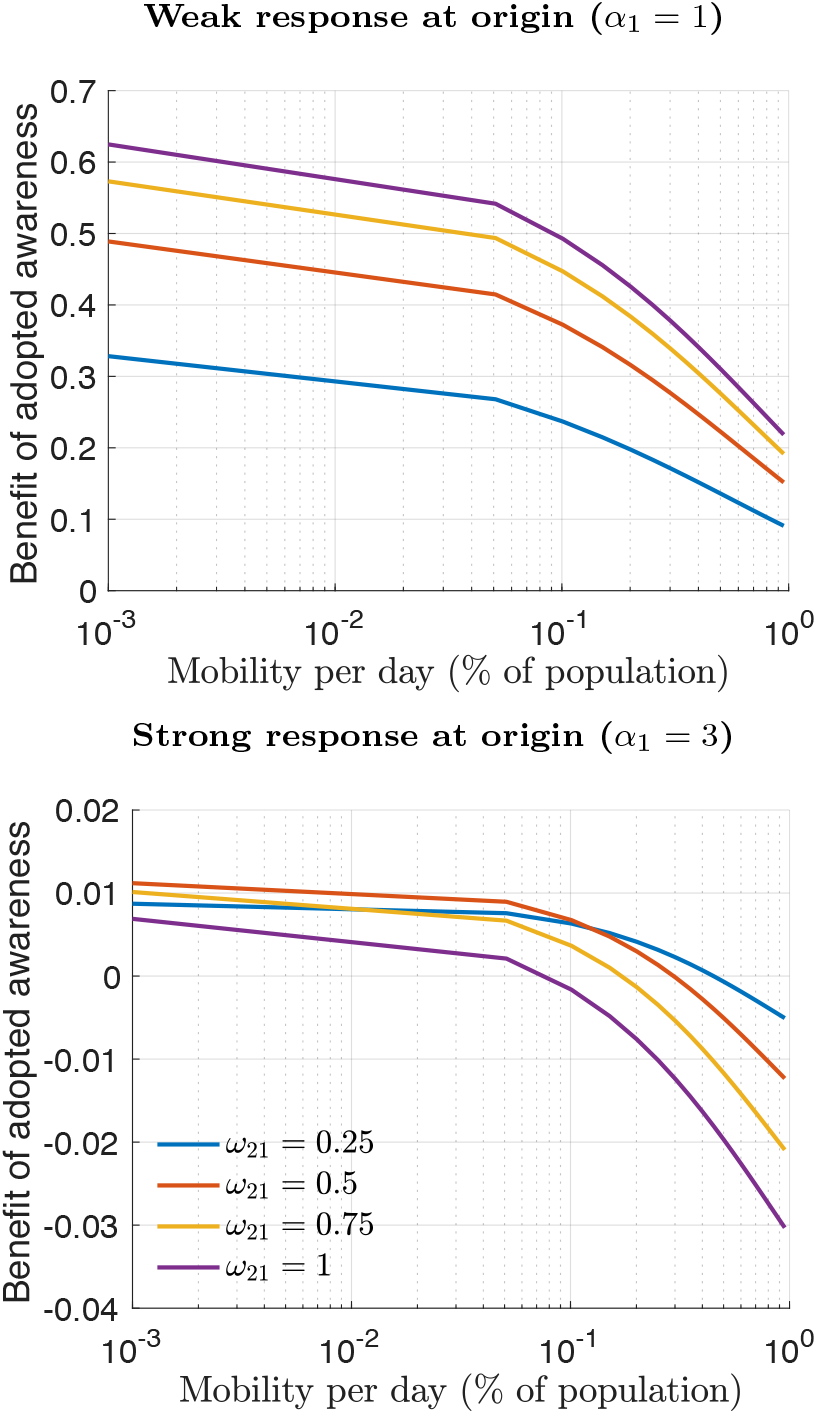
Benefit of adopted awareness with respect to mobility rates. (Top) Weak and (Bottom) strong responses at the origin. The strength of response at Locality 2 is weak *α*_2_ = 1. The benefit is measured as the reduction in final size with respect to the zero-adopted awareness constant scenario *ω*_21_ = 0. Let *F*_2_(*ω*_21_, *λ*_12_) denote the final outbreak size at Locality 2 with respect to *ω*_21_ and *λ*_12_. The benefit of alertness is defined as *F*_2_(0, *λ*_12_) − *F*_2_(*ω*_21_, *λ*_12_). Weak response at the origin, higher adopted awareness leads a smaller outbreak size at Locality 2 (Top). Given a strong response at the origin, higher adopted awareness can lead to higher outbreak sizes (Bottom). Strong response at the origin reduces the magnitude of the benefit of adopted awareness.

Thus far, we assumed the total flow of individuals from one locality to another is fixed and does not depend on the severity of the outbreak. An alternative is to let mobility rates be dependent on awareness. Flow to a locality that is experiencing a severe outbreak can reduce, and similarly flow from localities with widespread outbreaks toward regions with less severe outbreaks can increase—see Appendix C for one such awareness driven mobility dynamics. Another alternative is to reduce the overall flow to and from locality based on the current size of the outbreak. In a two locality setting where one locality is the origin, such awareness-driven mobility dynamics delay the time disease takes-off in the neighboring locality, increasing the time for neighboring locality to be better prepared. In turn, the benefit of awareness increases similar to the effect of reduction in mobility rates discussed above.

### 3.6. Effects of population sizes

We consider scenarios where the two localities have differing population sizes *N*_1_ ≠ *N*_2_. In the model dynamics given in (1)-(5), we assume the populations mix at a fixed rate *λ*_*ij*_. Thus, the population size differences would not affect the flow implying that former results would continue to hold even when *N*_1_ ≠ *N*_2_. We consider an alternative mobility model where the mobility constants *λ*_*ij*_ represent the flow rates in order to analyze the effects of population size differences—see Appendix C. In this alternative model, the amount of flow from one locality to another depends on the size of the originating compartment (*S*_1_, *E*_1_, *S*_2_, or *E*_2_) This model provides identical results when *N*_1_ = *N*_2_. When the initial population sizes are different (*N*_1_ ≠ *N*_2_), the mobility dynamics will generate flows such that the population sizes will change over time. In turn, this will affect the ratio of the cumulative infected *R*_2_(∞)*/N*_2_(∞) where we note that *N*_2_(∞) represents the size of the population in Locality 2 at time *t* =∞.

The effect of population differences is negligible when mobility is low (Fig. S4). The small differences in outbreak sizes are caused by the change in population sizes. For instance, when the population size at Locality 1 is larger than Locality 2 (*N*_1_ > *N*_2_), individuals flow from Locality 1 to Locality 2 increasing the population size of Locality 2. In turn, the fraction of recovered individuals in Locality 2 gets lower because its final population size is larger *N*_2_(∞) > *N*_2_. A secondary effect of different population sizes manifests when the origin goes through a worse outbreak and the importing locality is prepared. In this case, there is a larger migration of susceptible individuals from Locality 2 to Locality 1. This magnifies the outbreak ratio in Locality 2. All of the aforementioned effects are more pronounced when mobility rate is higher.

## 4. Conclusions

We developed a mathematical model to analyze the impact of social distancing efforts on disease dynamics among interconnected populations. We assumed that social distancing efforts at a given location is a function of both disease prevalence within the population and outbreak dynamics at neighboring localities. The inclusion of influence of outbreak size at neighboring localities distinguishes the model considered here from existing metapopulation models that only consider social distancing based on local disease prevalence [22, 15]. Our analysis showed that it is beneficial to reduce travel between localities given the inability to detect latently infected individuals (consistent with earlier findings [1]). However, this benefit is contingent on how prepared neighboring localities are for the importation of cases. We used the term adopted awareness to determine the importance given to preparedness at neighboring localities. We assumed the preparedness at importing localities is an increasing function of the outbreak size at the origin. The increasing function assumption implied that neighboring localities increase their levels of preparedness as the severity of the disease at the origin increased. That is, the severity of the outbreak at the origin triggers social distancing efforts at neighboring localities by local authorities making non-pharmaceutical interventions, e.g., declaring state of emergency or issuing stay at home orders. We derived an upper bound on the outbreak size at importing localities as a function of the outbreak size at the origin and strength of response at the importing locality. Using this upper bound, we identified a critical threshold for the adopted awareness weight that would eliminate the disease at importing localities.

It is not surprising that increased levels of preparedness reduces the outbreak size at localities neighboring the origin. However, the level of preparedness is dependent on the outbreak size at the origin. Thus, levels of preparedness increase at a locality when a neighboring locality has a larger outbreak size. Our results show that increased levels of preparedness at neighboring localities can yield lower total outbreak sizes even when the response at the origin is weak (Figure 5(Top)). The theoretical and numerical results mentioned above hold under a low mobility regime in which there is a lead time for increased alertness levels at importing localities based on the outbreak size at the origin. We identified that when the response at the origin is strong, adopted awareness may hurt, rather than benefit, the neighboring localities under higher mobility rates (Figure 6 (Bottom)). The neighboring localities may have, in effect, a false sense of security. In contrast, weak responses at the origin can paradoxically benefit neighboring localities that adjust their distancing based on adopted awareness. We also find that the benefit of adopted awareness is robust to small variations in mobility, awareness-driven mobility dynamics (Appendix C), heterogeneous population sizes (Section 3.6) and to variation in the inherent infection rate of the disease (Appendix D).

Overall, our findings imply that if there are multiple localities with outbreaks, the jurisdictions with less severe outbreaks should be looking at their worse-off neighbor rather than their best-off neighbor, and implementing social distancing measures accordingly. This finding provides further support for related work showing that coordination of responses can stop outbreaks when discordant responses do not [17]. The effects of awareness-driven social distancing and disease preparedness of connected communities during an epidemic outbreak should be further assessed using epidemiological models that account for important biological features of the disease. For instance, experiments on temporal viral shedding of COVID-19 estimate nearly half of the secondary cases happen by being in contact with individuals in pre-symptomatic stage [36]—see [4, 37] for other analysis of the impact of asymptomatic spreading. Here, we do not make a distinction between symptomatic and asymptomatic infected individuals; further extensions could incorporate such differences, e.g., [1, 37]. In addition, the current model does not account for the mobility of recovered individuals and their impact on reducing transmission, given disease-specific modification of behavior [30]. Such holistic approaches to modeling that include mechanistic social distancing terms in complex epidemiological models can provide an essential perspective on effective control of the pandemic [32]. This paper takes a step in this direction by providing analytical and numerical results on the importance of awareness-driven behavior and preparedness, and mobility.

## Data Availability

The code is available on Github page of the corresponding author.

https://github.com/ceyhuneksin/reacting_outbreaks_neighboring_localities

## Appendix A. A lower bound on the outbreak size at the origin

We derive a closed form solution for the outbreak size at the origin for the SEIR model in (1)-(4) when mobility is not included (*λ*_*ij*_ = 0). We modify the social distancing model at the origin (Locality 1) by

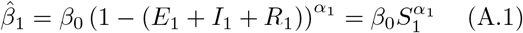

where we assumed *N*_1_ = 1 to simplify notation. The social distancing model above assumes that individuals distance with respect to the cumulative number of cases including the exposed individuals which were not included in (5). We note that this assumption is for analysis purposes only and allows us to compute a lower bound on the outbreak size for the original model in (5).

We define the following quantity to be the weighted sum of exposed and infected individuals

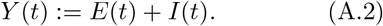

The force of infection is given by

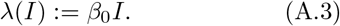

Given the force of infection and the reproduction number 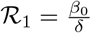, we can show that

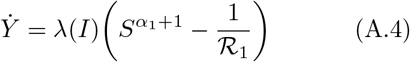

Then we have the constant of motion of the SEIR model ((1)-(4)) with the distancing model (A.1) as

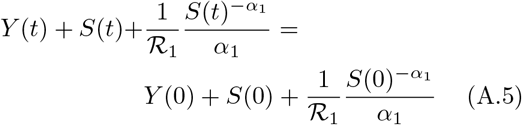

for any *t*. In identifying the above constant of motion, we divide *dY/dt* by *dS/dt* in (1), simplify terms, and integrate the resultant relation from time 0 to *t*. These steps are similar to the steps used to establish speed-outbreak size relations for standard SEIR models without social distancing, e.g., see [34, 35]. Now letting *t →* ∞ and using the fact that *S*(0) = 1, *Y* (0) = 0, *Y* (∞) = 0, we obtain the speed of spread versus final size relation in (6) for *α*_1_ > 0.

The social distancing function in (A.1) includes exposed individuals. That is, we have 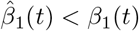 for all t where 1(t) is as defined in (5). Thus the final size R(∞) in (7) is a lower bound on the outbreak size at the origin.

## Appendix B. An upper bound for the outbreak size at the importing locality

Figures S1 and S2 show the lower bound on *S*(∞) obtained by solving (10). Top and bottom figures illustrate the change in the lower bound as a function of the strength of response at the origin. In accordance with the SEIR model (1)-(5), a strong response at the origin leads to a larger outbreak at Locality 2—compare diamond points in top and bottom panels in Figures S1 and S2. Similarly, the lower bound values 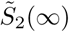 are lower in the bottom figures. Indeed, in both figures a weak response at the origin compounded by a strong response at Locality 2 guarantee that the disease does not spread in the Locality 2—see blue circles and diamonds in top figures.

**Figure S1:**
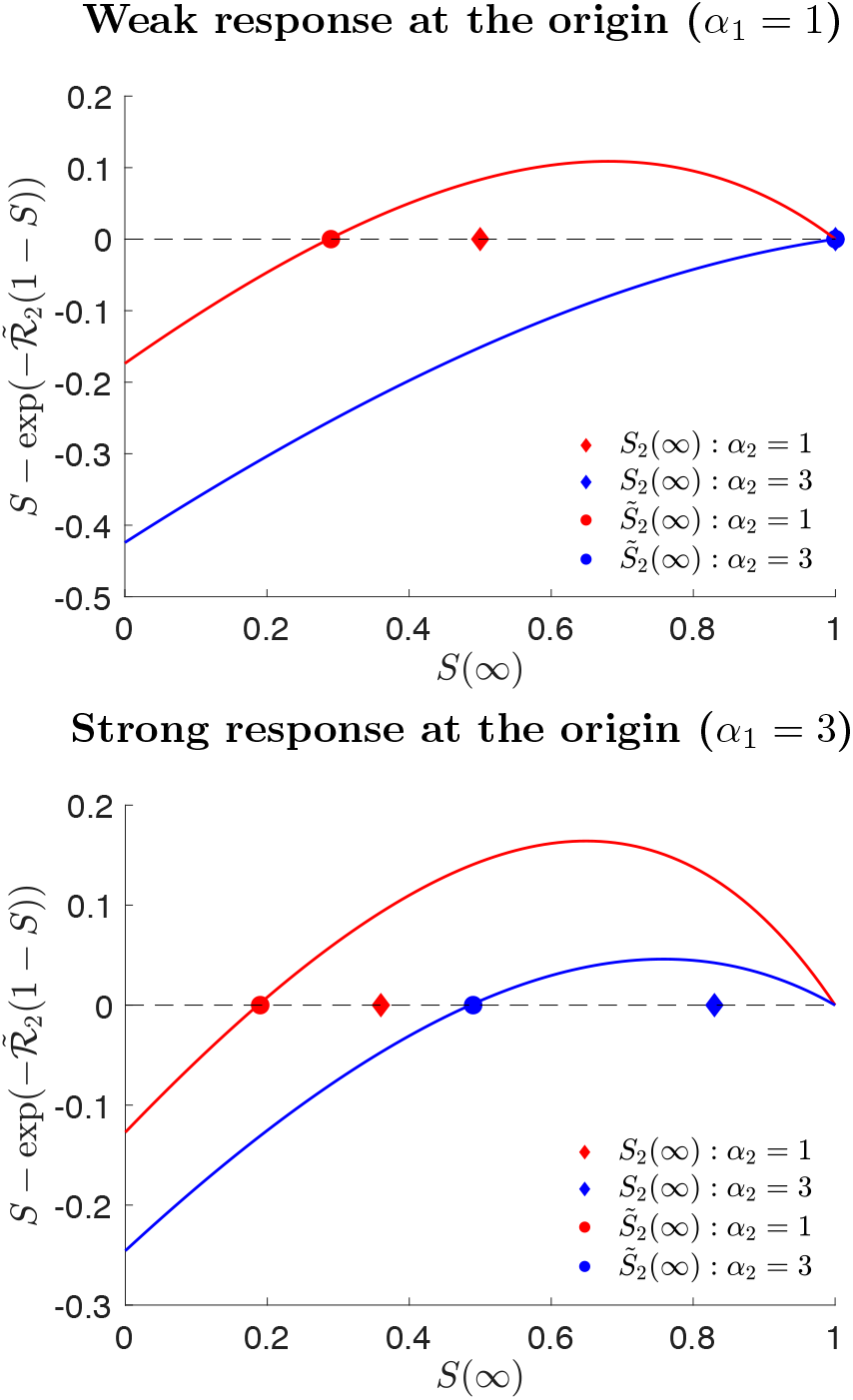
Lower bound values for *S*(∞). We assume low adopted awareness *ω*_21_ = 1*/*2. We let ℛ_1_ = 2.5 and *λ* = 0.0001%. (Top) Weak and (Bottom) strong response at the origin. Lines correspond to the left hand side of (10). Circle dots show the solution to (10), i.e., intersection of lines with zero. Diamond dots are *S*(∞) values obtained by simulating the SEIR model in (1)-(4) with *β*_*i*_ in (5).

The adopted awareness constant (*ω*_21_) is smaller in Figure S1 than in Figure S2. We observe that the lower bound for *S*(∞) is tighter when *ω*_22_ is smaller. This is expected since as *ω*_22_ decreases the importance given to prevalence at Locality 2 (*I*_2_ + *R*_2_) in (8). Thus the difference between the right hand sides of social distancing approximations in (8) and (9) decreases.

**Figure S2:**
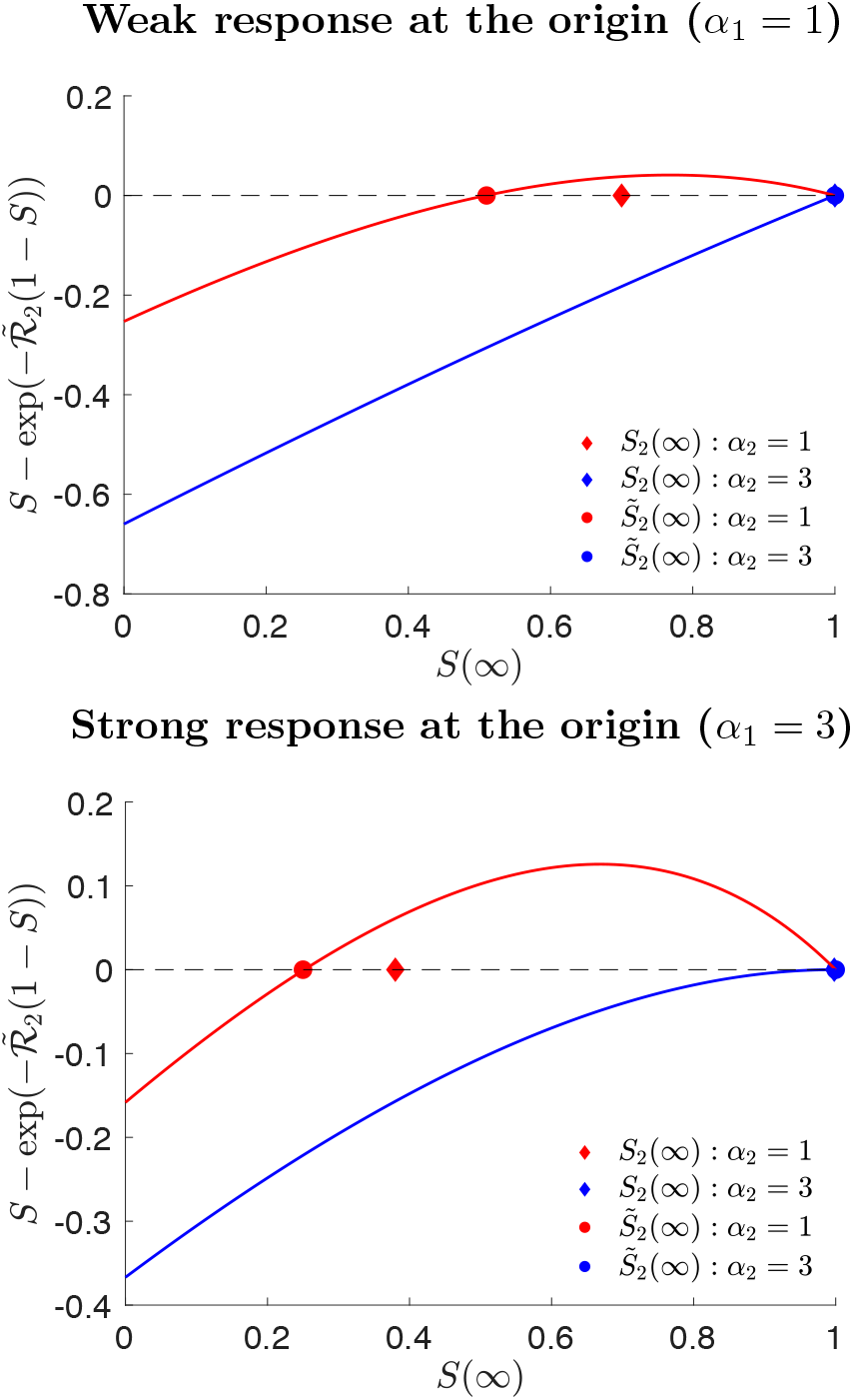
Lower bound values for *S*(∞). We assume high adopted awareness *ω*_21_ = 3*/*4. We let ℛ_1_ = 2.5 and *λ* = 0.0001%. (Top) Weak and (Bottom) strong response at the origin. Lines correspond to the left hand side of (10). Circle dots show the solution to (10), i.e., intersection of lines with zero. Diamond dots are *S*(∞) values obtained by simulating the SEIR model in (1)-(4) with *β*_*i*_ in (5).

## Appendix C. Alternative mobility dynamics

### Awareness-driven mobility dynamics

We consider a model where the flows between areas increase or decrease proportional to the ratio of current number of infected between localities, i.e.,

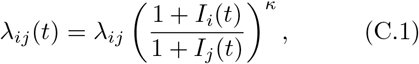

where *λ*_*ij*_ and *k* are positive constants. *k* determines the strength of mobility change as a function of the disease severity ratio. In the above model, the mobility flow from *i* to *j* increases as the ratio between the current outbreak size at locality *i* and locality *j* increases.

Fig. S3 shows how the ratio in (C.1) changes over time, indicating an increased flow at first from Locality 1 to Locality 2, and then an increased flow from Locality 2 to Locality 1 later. The difference in peak times of localities reduces as *k* increases. For instance, if *k* = 5, the reduction in the difference between peak times of localities ranges from 2% to 8% as adopted awareness constant *ω*_21_ increases from 0 to 1. This reduction in the difference between peak times do not lead to a meaningful change in the final outbreak sizes—see Fig. S3(Right).

### Population size dependent mobility dynamics

The modified model is as follows,

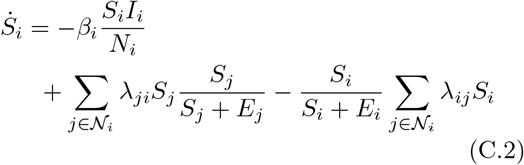

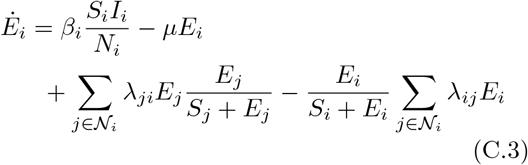

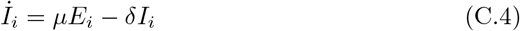

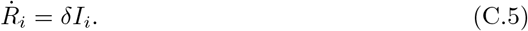

We note that the population sizes are changing over time when *N*_1_ ≠ *N*_2_ even if *λ*_*ij*_ = *λ*_*ji*_. We refer to the population size at locality *i* at time *t* using *N*_*i*_(*t*).

Given this modified model, we consider different population sizes for the origin (*N*_1_) with the ratio of the initial population sizes (*N*_1_*/N*_2_) ranging from 0.1 to 100 in Fig. S4. When the ratio (*N*_1_*/N*_2_) is smaller than 1, the model above yields a positive net flow from locality 2 to locality 1. When the ratio is larger than 1, the model above yields a positive net flow from locality 1 to locality 2.

## Appendix D. Sensitivity of benefit of awareness to variation in the infection rate

We consider the sensitivity of benefit of awareness with respect to variability in the inherent infection rate of the disease (*β*_0_). The infection rate affects the peak time and outbreak size at both localities which makes the direction of its effect on the benefit of awareness non-trivial. In particular, if infection rate *β*_0_ increases, the difference in peak times of two localities decreases, e.g., compare peak time difference values at *β*_0_ = 0.5 and *β*_0_ = 0.75 on the purple line in Fig S5. This creates less time for Locality 2 to prepare. At the same time, when the infection rate *β*_0_ is high, the outbreak size at the origin increases which leads to an increase in awareness at the Locality 2. We find that the latter effect slightly dominates the former effect yielding a minor increase in the benefit of awareness as the infection parameter *β*_0_ increases—see Fig. S5.

**Figure S3:**
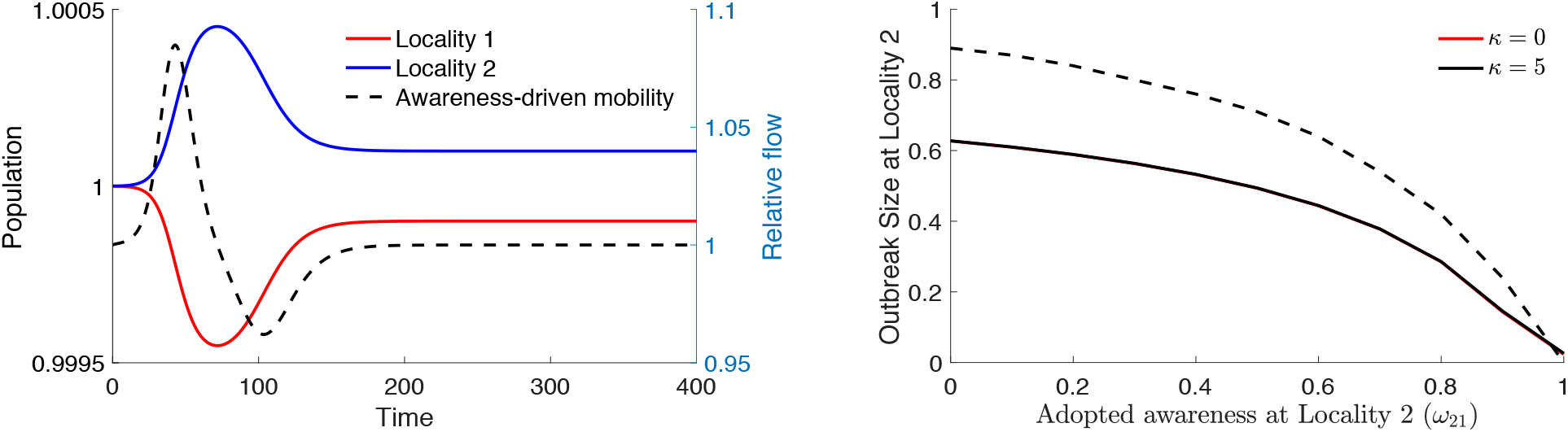
(Left) Population size and mobility flow changes over time. Left y-axis shows the population sizes of localities. Right y-axis shows relative mobility flow from locality 1 to locality 2 over time. (Right) Outbreak size at Locality 2. The flow *λ*_12_(*t*) is as given in (C.1) with *k* = 5. The rest of the parameters of the model (1)-(4) are *β*_0_ = 5*/*8, *µ* = 1*/*3, *δ* = 1*/*4, and *λ*_12_ = 0.001%, *N*_1_ = *N*_2_ = 1, *α*_1_ = *α*_2_ = 1 and *ω*_21_ = 0.5.

**Figure S4:**
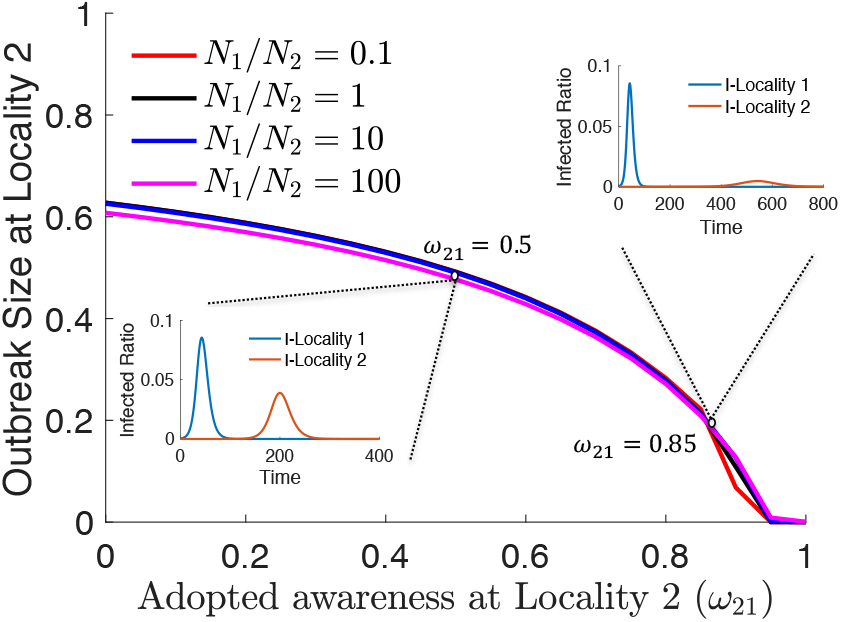
Outbreak size at locality 2 as a function of the population size ratio. Population size ratio is given by the ratio of initial population sizes between localities, i.e., *N*_1_*/N*_2_. The epidemic and mobility dynamics are given by (1)-(4) and eq. (5) in the revised manuscript. Parameters: *β*_0_ = 5*/*8, *µ* = 1*/*3, *δ* = 1*/*4, *λ* _12_ = *λ* _21_ = 10^−7^, and *α*_1_ = *α*_2_ = 1.

**Figure S5:**
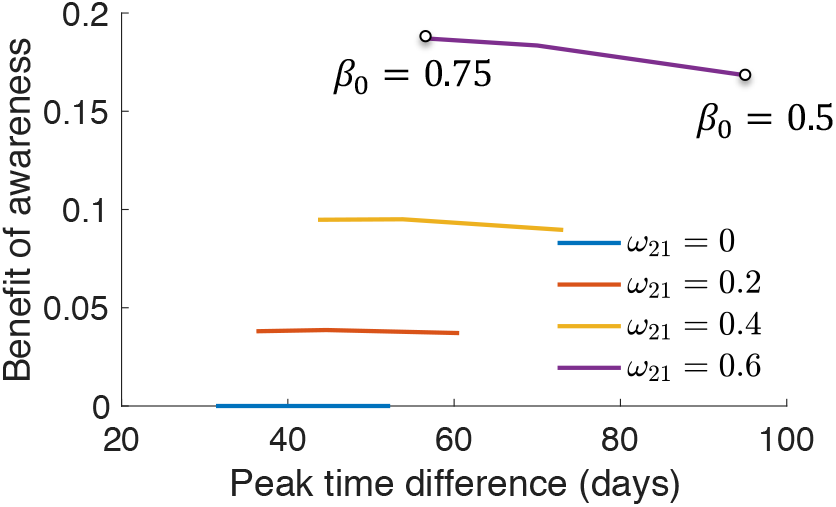
Benefit of adopted awareness versus the change in peak times as the infection rate changes. We let *β*_0_ ∈ [0.5, 0.75]. The peak time difference is the difference between the time Locality 2 peaks and the time Locality 1 peaks. Let *F*_2_(*ω*_21_, Δ*β*) denote the final outbreak size at Locality 2 with respect to *ω*_21_ and Δ*β*. The benefit of awareness is defined as *F*_2_(0, Δ*β*) − *F*_2_(*ω*_21_, Δ*β*) where Δ*β* ∈ [−0.125, 0.125]. Parameters of the dynamics (1)-(5): *µ* = 1*/*3, *δ* = 1*/*4, *N*_1_ = *N*_2_ = 1, *α*_1_ = *α*_2_ = 1, and *λ*_12_ = *λ*_21_ = 10^−5^.

## Acknowledgements

Ceyhun Eksin was, in part, supported by grants from the National Science Foundation (NSF CCF-2008855 and NSF ECCS-1953694). Martial-Ndeffo Mbah was, in part, supported by a grant from the National Science Foundation (DEB 2028632). Joshua S. Weitz was supported, in part, by a grant from the Army Research Office (W911NF1910384).

## Materials

The code is available at https://github.com/ceyhuneksin/reacting_outbreaks_neighboring_localities.

